# A Longitudinal Exploration of *CACNA1A*-related Hemiplegic Migraine in Children

**DOI:** 10.1101/2024.06.14.24308953

**Authors:** Donna Schaare, Laina Lusk, Alexis Karlin, Michael C. Kaufman, Jan Magielski, Sara M. Sarasua, Kendra Allison, Luigi Boccuto, Ingo Helbig

## Abstract

**Introduction:** Since the initial description of *CACNA1A-*related hemiplegic migraine (HM), the phenotypic spectrum has expanded from mild episodes in neurotypical individuals to potentially life-threatening events frequently seen in individuals with developmental and epileptic encephalopathies. However, the overall longitudinal course throughout childhood remains unknown.

**Methods:** We analyzed HM and seizure history in individuals with *CACNA1A*-related HM, delineating frequency and severity of events in monthly increments through a standardized approach. Combining these data with medication prescription information, we assessed the response of HM to different agents.

**Results:** Our cohort involved 15 individuals between 3 and 29 years (163 patient years) and included 11 unique and two recurrent variants (p.R1349Q and p.V1393M; both *n=*2). The age of first confirmed HM ranged from 14 months to 13 years (average 3 years). 25% of all HM events were severe (lasting >3 days) and 73% of individuals had at least 1 severe occurrence. Spacing of HM events ranged from 1 month to 14 years and changes in HM severity over time of showed increases or decreases of >2 severity levels in 12/122 events. Eight individuals had epilepsy, but severity of epilepsy did not correlate with frequency and severity of HM events. While levetiracetam (*n=*6) and acetazolamide (*n=*5) were the most frequently used medications, they did not show efficacy in HM prevention or HM severity reduction. However, verapamil (*n=*3) showed efficacy in preventing HM episodes (OR 2.68, CI 1.39-5.67).

**Significance:** The longitudinal course of *CACNA1A*-related HM lacks recognizable patterns for timing and severity of HM events or correlation with seizure patterns. Our data underscores the unpredictability of *CACNA1A*-related HM, highlighting the need for close surveillance for reoccurring HM events even in individuals with symptom-free periods.

**Key points:**

1. 24% of hemiplegic migraines (HM) in *CACNA1A-*related disorders are severe, involving cerebral edema and greater than 4 days to recover
2. Timing and severity of HM are unpredictable, with large changes in severity between events, and age of onset ranging from 1-13 years
3. Epilepsy occurred in 53% of individuals, with neither the timing nor severity of seizures correlated with HM

## Introduction

Hemiplegic migraine (HM) is a rare neurological condition associated with pathogenic variants in several identified genes, primarily *CACNA1A* and *ATP1A2*, and less frequently *SCN1A*, *PRRT2,* and *SCN2A.*^1–4^ Initially described in 1996 as a rare subtype of migraine with aura, HM is presently defined by the International Classification of Headache Disorders (ICHD-3) as a phenomenon characterized by transient unilateral motor weakness during the aura phase lasting minutes to weeks with additional symptoms including visual, sensory, and dysphasic abnormalities; accompanying headache is not necessary for the diagnosis.^5,6^ HM caused by pathogenic variants in *CACNA1A* (located on chromosome 19p13.13) have historically been classified as either Familial Hemiplegic Migraine type 1 (FHM1), resulting from inherited variants, or Sporadic Hemiplegic Migraine type 1 (SHM1), resulting from *de novo* pathogenic variants. The literature has suggested a correlation between severity and underlying genetic etiologies, not only in the manifestations of the hemiplegic events but in the overarching phenotype of the patients, including baseline neurodevelopmental trajectory.^1,2,7–10^ However, these classifications prove inadequate in wholly describing the phenotype of individual patients. In particular, these definitions do not address the more severe presentations of HM that can be seen within the pediatric population, which can range from mild episodes of transient weakness to severe episodes of protracted hemiplegia, encephalopathy, seizure, and even life-threatening cerebral edema.^9^ Consequently, they underestimate the potential severity and urgency of events in this population.

*CACNA1A-*related HM is one of several *CACNA1A*-related neurological conditions, which also include epilepsy, neurodevelopmental disorders, and episodic and progressive ataxia, among other movement disorders.^1,2,7–10^ Prior studies, including case reports, have described the clinical features of HM events in individuals with pathogenic *CACNA1A* variants, but these have rarely conducted a longitudinal analysis of event frequency and severity.^4,11–15^ In addition, while these case reports have narratively reconstructed patient histories, they have rarely surpassed two to three individuals per report.^1,2,4,7,8,10,11,13,15–26^ In contrast, most studies with relatively large sample sizes have reported on overall lifetime features rather than evolution of features over time or trends in events over the lifetime, and have reported features independently of one another (i.e. looking at seizures and HM independently without examining the possible bidirectional relationships between the occurrences of either).^11,15,19,27^ Finally, data on treatment efficacy for *CACNA1A*-related HM is largely limited to anecdotal accounts of single cases or small numbers of patients.^4^ Taken together, these factors render *CACNA1A*-related HM an elusive phenomenon with an unclear clinical pattern and unexplored trajectories.

While prospective studies pose significant challenges for rare genetic conditions such as *CACNA1A*-related HM, electronic medical record (EMR) data can be used to comprehensively delineate clinical spectra of such genetic neurodevelopmental disorders and epilepsies.^28–30^ Human Phenotype Ontology (HPO), a standardized biomedical ontology, serves as a powerful tool to standardize the wealth of heterogeneous clinical information and allow high-throughput analyses on it.^31–33^ By employing a systematic computational approach to information contained within the EMR, data can be successfully harmonized and tracked over time, allowing the in-depth characterization of a given condition and relevant clinical subgroups within it.

Here, leveraging the HPO framework and standardized computational approaches to the EMR data, we investigate the complexity of *CACNA1A*-related HM within the pediatric population by delineating the longitudinal time course of HM events and seizure history in 15 unrelated individuals with known pathogenic or likely pathogenic *de novo* variants in *CACNA1A* and a diagnosis of at least one lifetime HM. We describe the incidence and severity of HM events, comorbid epilepsy, neurodevelopmental phenotypes, and medication histories. This longitudinal cohort analysis contributes to a more complete phenotypic characterization of *CACNA1A*-related conditions in a pediatric population.

## Materials & Methods

### Identification of individuals with *CACNA1A*-related HM

The cohort for this longitudinal analysis was obtained from individuals enrolled in the Epilepsy Genetics Research Project (EGRP) at the Children’s Hospital of Philadelphia (CHOP, Philadelphia, PA, USA) and patients enrolled in a similar study from the Cleveland Clinic (CCF, Cleveland, OH, USA). The international cohort included individuals from both the United States and Australia. Inclusion criteria included documentation of a pathogenic or likely pathogenic variant in the *CACNA1A* gene and at least one lifetime HM. Informed consent was obtained from the parents of all participants in agreement with the Declaration of Helsinki, and the study was performed following local protocols approved by the institutional review boards (IRBs) at CHOP or CCF. Of note, two individuals were previously reported as case reports (Individual #5^14^ and Individual #14^34^). Clinical information was obtained via medical records accessed by a clinician or the family. *CACNA1A* variants were identified through prior clinical testing and involved whole exome sequencing (*n*=8), epilepsy panel (*n*=3), single gene testing (*n*=2), HM panel (*n*=1), or autism panel (*n*=1).

### Assigning Human Phenotype Ontology terms to phenotypic manifestations

We manually extracted the phenotypic manifestations from the electronic medical records (EMRs) and chart notes and assigned HPO terms (https://hpo.jax.org/app/, accessed in October 2023). Additionally, in collaboration with clinical experts, we created an ontology for features observed during HM episodes compatible with the HPO framework in order to harmonize all phenotypes present in *CACNA1A*-related disorders. Automatic reasoning was performed to assign higher-level HPO terms by identifying an initial “base” term and a “propagated” term to overcome the heterogeneity of the depth of the HPO terms as has been previously described.^28,30,35^

### Reconstruction of hemiplegic migraine event history in monthly intervals

To recreate the HM history of each study participant, we reviewed the medical records for the occurrence of HM. We recorded the frequency and severity of events in monthly increments, employing the analogical strategy as with the reconstruction of seizure histories our group performed in the past.^28^ We defined an HM event as per the ICHD-3 for consistency.^5^ We recorded each patient’s neurological history, including the age of the first diagnosed HM, any preceding episodes of concern, where relevant, and the age of the last assessment. Once identified, the severity for each event was graded on an increasing ordinal scale from Grade 1 to 5 (based on time to recovery to baseline and duration of hospitalization) along with associated clinical features of the event, where features such as cerebral edema, encephalopathy, reduced consciousness, status epilepticus, and abnormal MRI/EEG were associated with a higher score (see **Table 1**). Grade 1 was transient and required no visit to the Emergency Department (ED). Grade 2 could involve an ED visit but did not require a transfer to the Intensive Care Unit (ICU). Grade 3 had a longer duration, 1-3 days, and could involve admission to the ICU. Grade 4 involved more than 3 days of hospitalization before return to baseline but less than 1 week. Grade 5 events required more than 7 days of hospitalization and recovery time. Severe features were isolated to Grade 4 and Grade 5 events which correlated reasonably well with time to recovery. The age of the first documented abnormal brain imaging on either MRI and/or CT was also recorded.

**Table 1.**
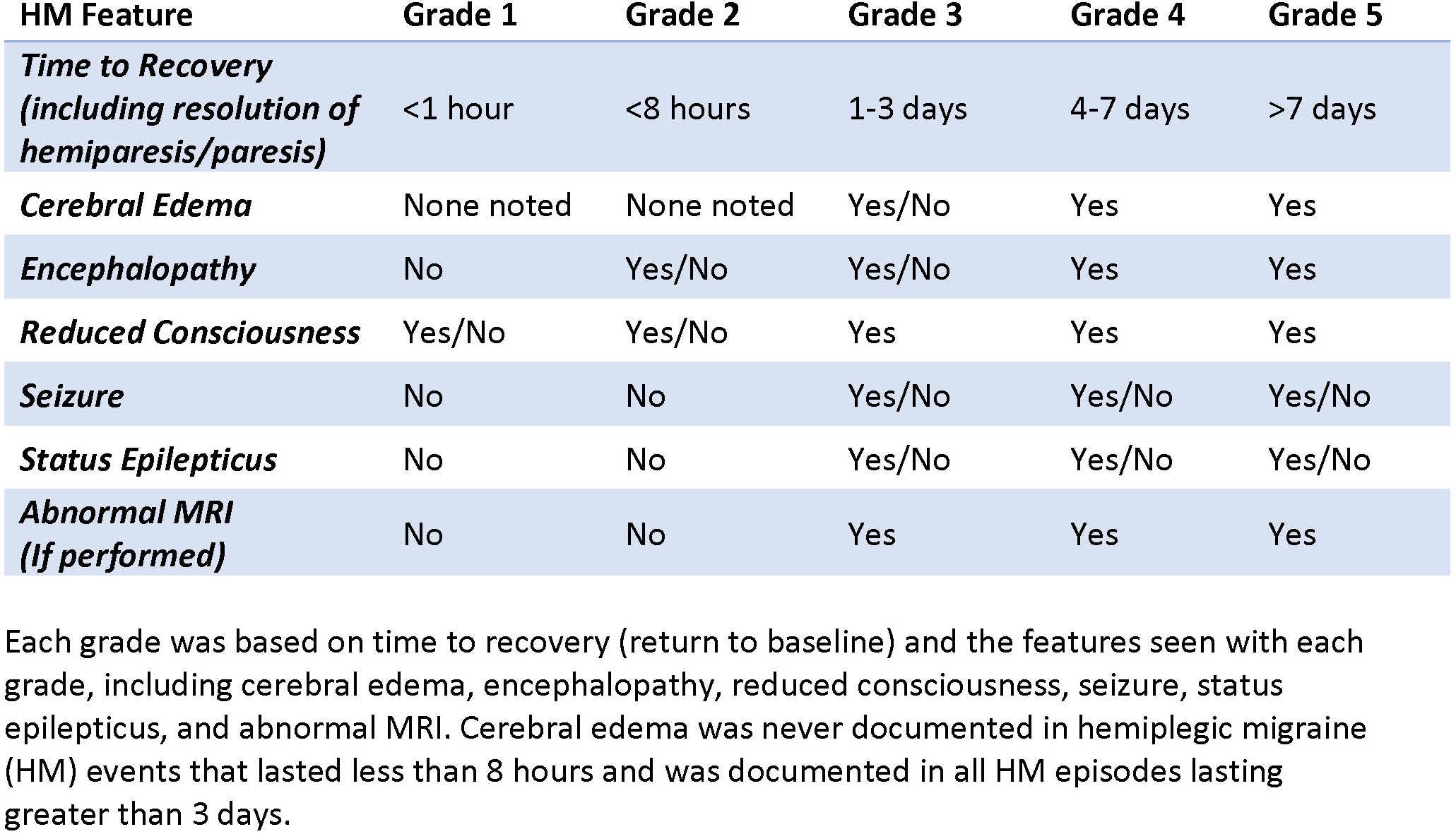
Clinical features associated with hemiplegic migraine severity grades.

### Reconstruction of seizure history in monthly increments

In individuals with seizures, we also recorded the frequency and severity of seizures in monthly increments. We used HPO terms to label the seizure types. We employed the protocol by Xian et al. to assign seizure severity and plotted the frequency over time for the life of each individual^28^. To summarize, seizure severity was defined on an increasing scale from 0 to 5 and correlated to the number of seizures in a given month (0 = none, 1 = monthly, 2 = weekly, 3 = daily, 4 = 2-5 per day, 5 = >5 per day).

### Delineation of preventative therapeutics for hemiplegic migraine

We reviewed medical records to document the use of preventive therapies for HM and/or epilepsy over time for each study participant. Similarly to how our group assessed the effectiveness of anti-seizure medications in the past, we retrospectively assessed the comparative effectiveness of preventing HM by examining the frequency and severity of HM episodes compared with the prior month to determine if there was an increase or decrease in HM frequency or severity.^28,36^ Statistical significance of the odds ratios were calculated with the use of a 95% Confidence interval and the use of Fisher’s exact test.

### Statistical analysis

All computations were performed using the R Statistical Framework.^37–39^ Statistical testing for associations is reported with correction for multiple comparisons using False Discovery Rate (FDR) of 5%. In cases where statistical significance was not reached after correction for multiple comparisons, findings remain descriptive and are presented as odds ratios with 95% confidence intervals. Primary data for this analysis is available in the supplementary material. Computer code for all analysis is available on request.

## Results

### Demographic and genetic information

The 15 individuals with confirmed *CACNA1A*-related HM included eight males (53%) and seven females (47%), ranging in age at time of data collection from 3 to 29 years old. Age at diagnosis ranged from 1 year to 10 years. Phenotypic features and variant details are listed in **Table 2**. Fourteen individuals had missense mutations in either the S4 voltage sensor or the S5 and S6 pore transmembrane regions of the protein (**Figure S1**). One individual had a missense variant located in the intracellular loop between S5 and S6 (**Figure S1**, Individual #2). Variants were distributed across all 4 protein domains.

**Table 2.**
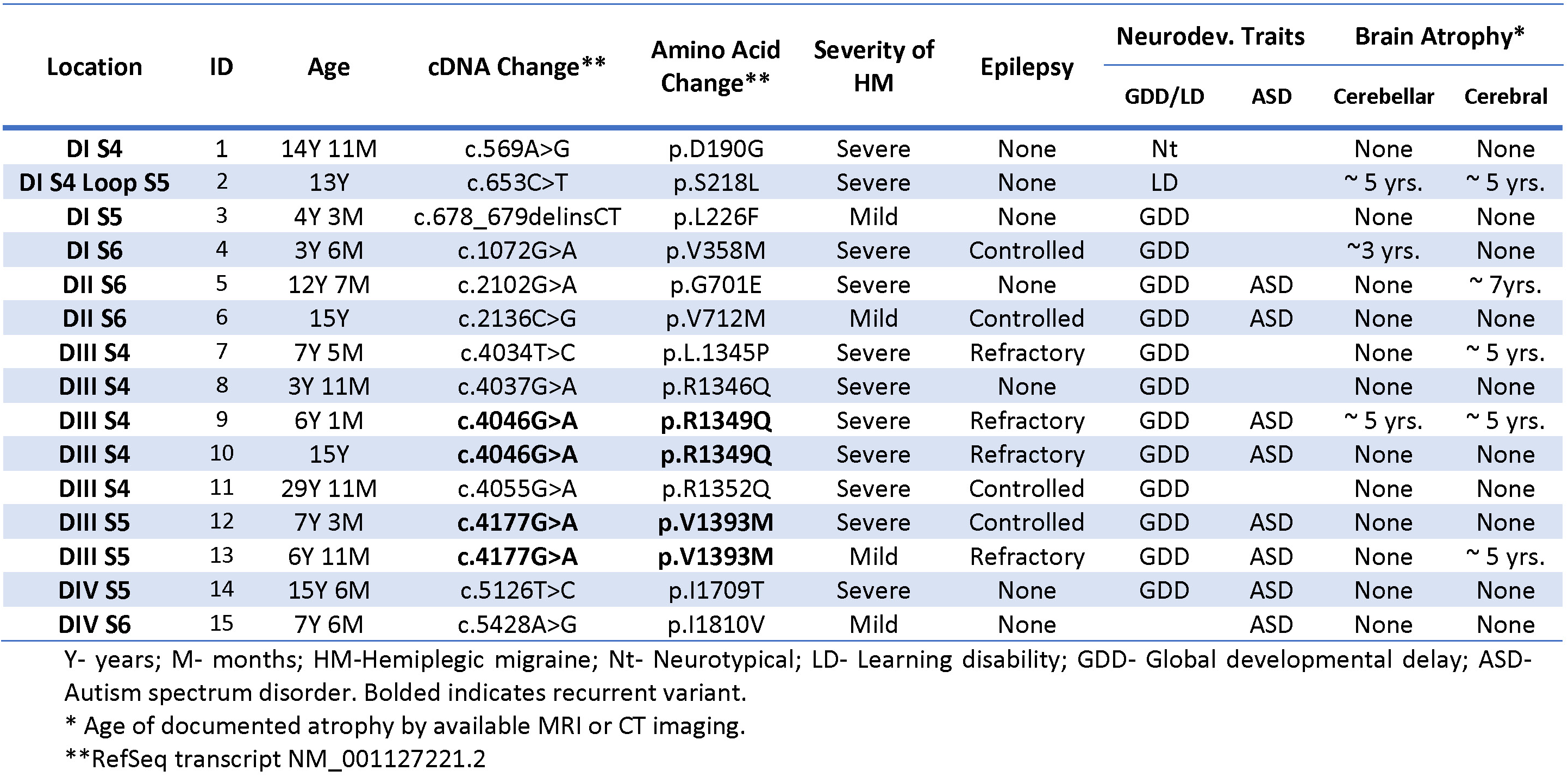
Genotypes and Corresponding Phenotypes in this Cohort.

### The phenotypic spectrum in *CACNA1A*-related HM demonstrates broad variability

Seventy-seven clinical concepts in 12 categories (coded in HPO terminology) were assigned to the cohort (**Table S1**). These terms accounted for clinical presentations during both the overall disease trajectory and HM events specifically. Phenotypic features outside of HM episodes included neurodevelopmental features (**Figure 1A**), seizures, and other features that appear either chronically or episodically.

**Figure 1.**
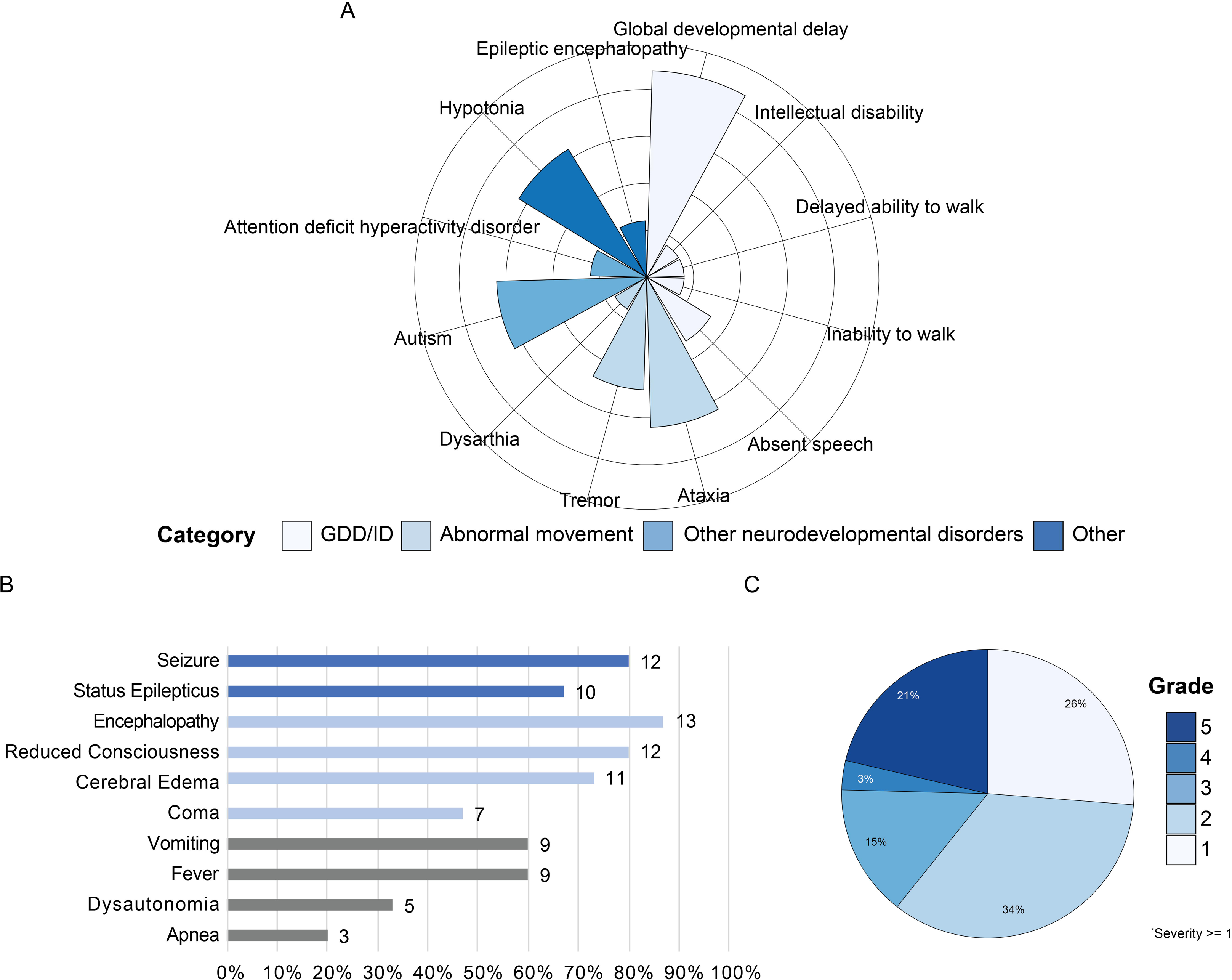
Phenotypic features of the cohort. **(A)** Atypical neurodevelopmental traits, categorized by color: white - global developmental delay (GDD)/intellectual disability (ID) and related terms; light blue – abnormal movements; blue – other neurodevelopmental disorders; dark blue - other. Numbers next to bars indicate the number of individuals (out of 15 total). **(B)** Manifestations observed during hemiplegic migraine episodes. Features grouped by color: dark blue - seizure-related features; light blue - other severe features; dark gray - other non-severe features. **(C)** Distribution of hemiplegic migraine severity scores for all hemiplegic migraine events reported in the cohort. The most frequent hemiplegic migraine severity was Grade 2 at 34.4%, followed by Grade 1 at 26.2%. Severe episodes (Grades 4 and 5) accounted for 25% of all episodes.

#### Hemiplegic migraine episodes

In total, there were 122 episodes across 15 individuals. During HM events, 13/15 (86.7%) individuals had at least one episode associated with encephalopathy, 12/15 (80.0%) with reduced consciousness, 12/15 (80.0%) with seizures, 11/15 (73.3%) with documented cerebral edema, 10/15 (67.7%) with status epilepticus, 9/15 (60.0%) with fever, and 9/15 (60.0%) with vomiting (**Figure 1B**). Less common features included coma (7/15, 46.7%), dysautonomia (5/15, 33.3%), and apnea (3/15, 20.0%). The duration of HM episodes ranged from less than 30 minutes (Grade 1, n=32/122 total episodes, 26.2%), to greater than 7 days (Grade 5, n=26/122 total episodes, 21.3%). The most common duration was greater than 30 minutes but less than 8 hours (Grade 2, n=42/122 total episodes, 34.4%). Durations of 1-3 days (Grade 3, n=18/122 total episodes, 14.8%) and 4-7 (Grade 4, n=4/122 total episodes, 3.3%) were less common **Figure 1C**). Overall, 11/15 (73.3%) individuals experienced at least one HM that would be considered severe (involving cerebral edema or lasting more than 3 days, corresponding to grades 4 or 5. In total, approximately 25% of the HM events were severe while the majority of HM episodes were of mild severity (lasting 3 days or less, corresponding to Grades 1-3). Although Grade 3 HM events could involve edema, it was usually mild, unlike the edema seen in Grades 4 and 5, correlating to the shorter time to return to baseline. Grade 4 (resolving within 4-7 days) was the least common, representing less than 5% of all HM episodes, suggesting that once an HM episode surpasses 4 days, it is likely to last for a minimum of a week and involve significant cerebral edema (**Figure 1C**). Of note, 12/122 HM events involved a change of greater than 2 severity grades from the prior event (e.g. grade 3 HM followed by grade 5 HM). Six individuals reported minor head trauma as a trigger in at least one event and four patients cited infections as possible triggers.

#### Neurodevelopmental features

Outside of episodic events, we gathered data on neurodevelopmental features, coded into HPO terminology (**Table S1**). In our cohort, 12/15 (80%) individuals had neurodevelopmental abnormality, including global developmental delay (GDD) or intellectual disability (ID). At the time of data collection, 3/15 (20%) individuals were non-ambulatory and 4/15 (26.7%) were non-verbal with an additional 4/15 (26.7%) having communication challenges, including speech apraxia and dysarthria. Three (20.0%) individuals were diagnosed with epileptic encephalopathy and eight (53.3%) individuals had hypotonia. Over half of the cohort had abnormal movements including ataxia (8/15, 53.3%), tremor (5/15, 33.3%), paroxysmal tonic upgaze (PTU; 3/15, 20.0%), and nystagmus (6/15, 40.0%). Neuropsychiatric disorders were present and included autism (8/15, 53.3%), attention deficit hyperactivity disorder (ADHD; 3/15, 20.0%), anxiety (2/15, 13.3%), paranoia and suicidal ideation (1/15, 6.7%), and self-injurious behavior (1/15, 6.7%) (**Figure 1a**).

#### Seizures and epilepsy

Comorbid epilepsy was diagnosed in 8/15 (53.3%) individuals. The mean age of seizure onset was 22 months, and median age of onset was 12 months (range 3 months to 6 years). Focal seizures were the most common seizure type present in in 6/8 individuals with epilepsy (75%). Status epilepticus outside of HM episodes occurred repeatedly in a single individual. In addition, 4/8 individuals with epilepsy had seizures refractory to anti-seizure medications (50%). The most common abnormal interictal EEG findings were focal epileptic discharges (4/8 individuals with epilepsy, 50%) and abnormal slow frequencies (3/8 individuals with epilepsy, 37.5%). In the three individuals diagnosed with epileptic encephalopathy, seizure onset was at 3 months, 24 months, and 72 months, respectively.

#### Neuroimaging features

Cerebral atrophy was identified on imaging in 5/15 individuals (33.3%) and cerebellar atrophy was identified in 3/15 individuals (20%), both at a median age of 5 years. Two of the three individuals with cerebellar atrophy also had cerebral atrophy. Of those with documented atrophy, volume loss in the cerebellum was not apparent on imaging before the age of 3 or in the cerebrum before the age of 5 (**Table 2**).

### Longitudinal analysis highlights the range of severity and unpredictability of HM events

Frequency and severity of each HM were recorded monthly for each individual from birth through the age at data capture, totaling 1,956 patient-months (163 patient-years, **Figure 2**). First documented HM episode ranged from 14 months to 13 years, with a mean age of onset of approximately 4 years and a median age of 3 years. No events with hemiplegia or hemiparesis occurred before 1 year of age. The frequency and intensity of confirmed HM episodes varied for each individual, with no distinct pattern emerging. Four individuals (26.7%) never experienced severe episodes (Individuals #3, 6, 13, 15), while others (2/15, 13.3%) had the most severe type of event (grade 5) nearly annually (Individuals #5 and 9). Two individuals (13.3%) had a later onset of HM episodes, experiencing their first HM event at age 10 and 13, respectively (Individuals #1 and 14). Others experienced more frequent attacks that began at a much younger age (5/15, 33.3% experienced > 3 HM events under 5 years of age, Individuals #3, 5, 6, 11, and 13).

**Figure 2.**
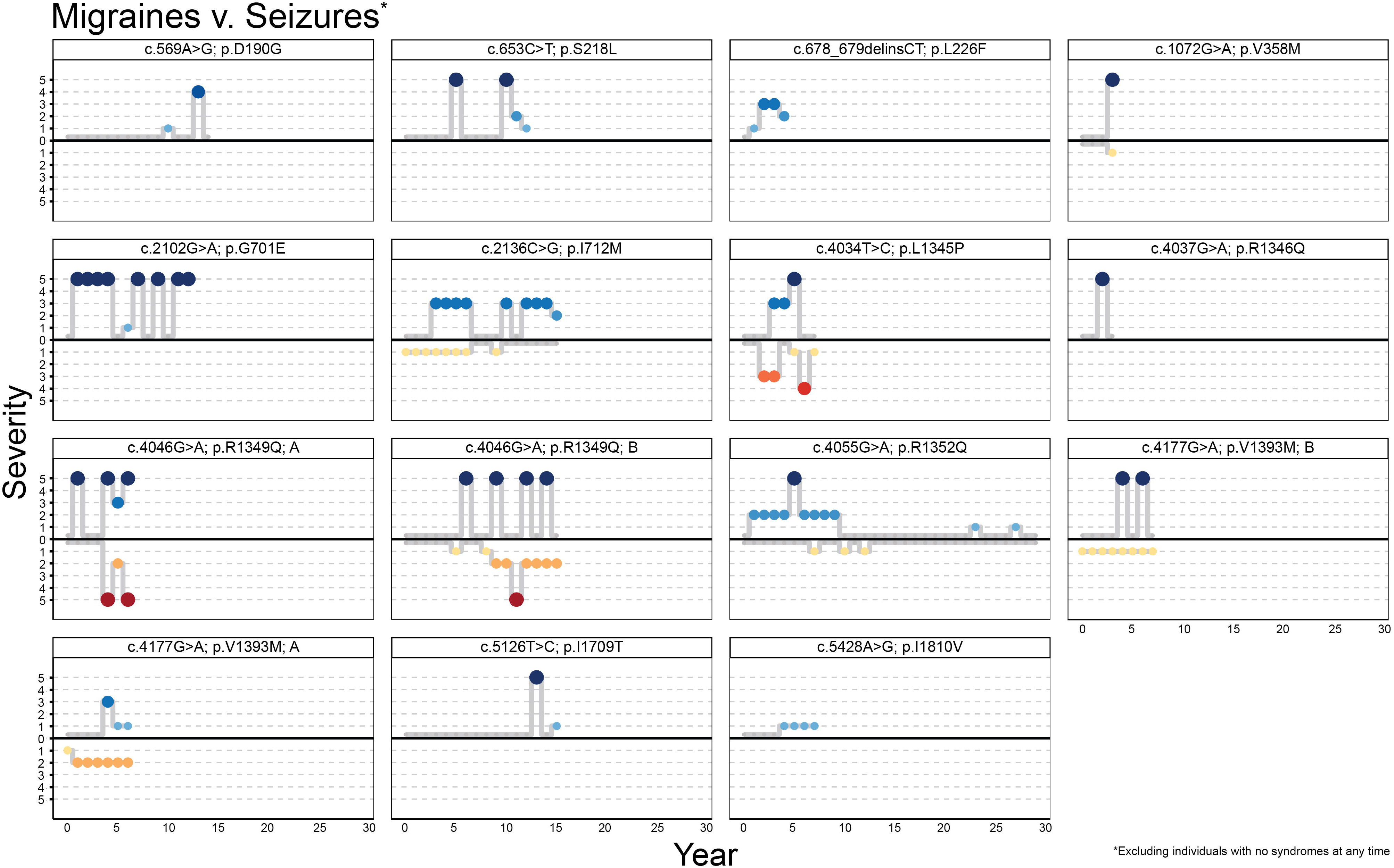
Longitudinal analysis of hemiplegic migraine and seizure events per year for each individual in the cohort. All events are plotted left to right, with graphs starting at age 0. (Note: Multiple events within a year were collapsed into one event) Intensity is ranked from 1 (mildest-smallest dot and lightest color) to 5 (most severe-largest dot and darkest color). Above the x-axis: hemiplegic migraine (HM) events (shades of blue). Below the x-axis: Seizures (shades of yellow to red). Each individual demonstrated their own unique pattern. A noteworthy finding was that no HM events occurred before the age of one.

For 4/15 (26.7%) of the individuals in the cohort, episodes suspicious for encephalitis without hemiplegia occurred before the first confirmed HM. These events involved altered consciousness, fever, and often status epilepticus. Individual #13 experienced the earliest of such episodes at 3 months and then again at 5 and 6 months with status epilepticus occurring each time. Individual #6 experienced six events after mild head trauma beginning at age 8 months and included status epilepticus, altered consciousness with and without fever. Individual #12 experienced an event at 9 months with global weakness that lasted approximately 2 hours. Finally, Individual #5 was the oldest to experience these types of events, which occurred at 22 months with status epilepticus and 25 months with global weakness that resolved in less than an hour. The duration of these episodes ranged from a few hours to greater than 3 days. Given that the diagnostic work-up in these episodes was negative, it may be reasoned that these episodes represent early HM events.

### Seizures have variable severity over time and weakly associate with HM events

As with HM presentation, the seizure severity varied within and between individuals in the cohort. Seizure frequencies ranged from monthly (severity score 1) to multiple times per day (severity score 5). The correlation between the severity of epileptic activity and simultaneous HM events associated with intensity was weakly associated with a marginal R^2^ value of 0.37. **Figure S2** is a visual depiction of the association but not a reflection of the correlation analysis.

### Preventative medication data suggests drug efficacy in reducing HM frequency or recovery time to baseline, but not both

Medications primarily used to prevent future HM events were used in 10/15 (66.7%) individuals within the cohort and included verapamil, acetazolamide, and topiramate (**Figure 3**). Additionally, 14/15 (93.3%) were managed with antiseizure medications, including valproate, lacosamide, oxcarbazepine, and clobazam. Of these, 8/14 (53.3%) were diagnosed with epilepsy, and as noted earlier, 4/8 (50%) individuals with epilepsy were refractory to two or more medications. Outside of medications, epilepsy treatments included a partial left temporal lobectomy in one individual who ultimately became seizure free (Individual #6). Two individuals had vagal nerve stimulator (VNS) placement and two individuals were on the ketogenic diet at some point during the disease trajectory. Neither VNS placement nor the ketogenic diet had clear effects on reducing HM frequency or severity. Abortive medications for HM events applied in our cohort included steroids and acetazolamide. The ability to analyze the benefit of abortive treatments was limited due to a small number of events when such strategies were applied.

**Figure 3.**
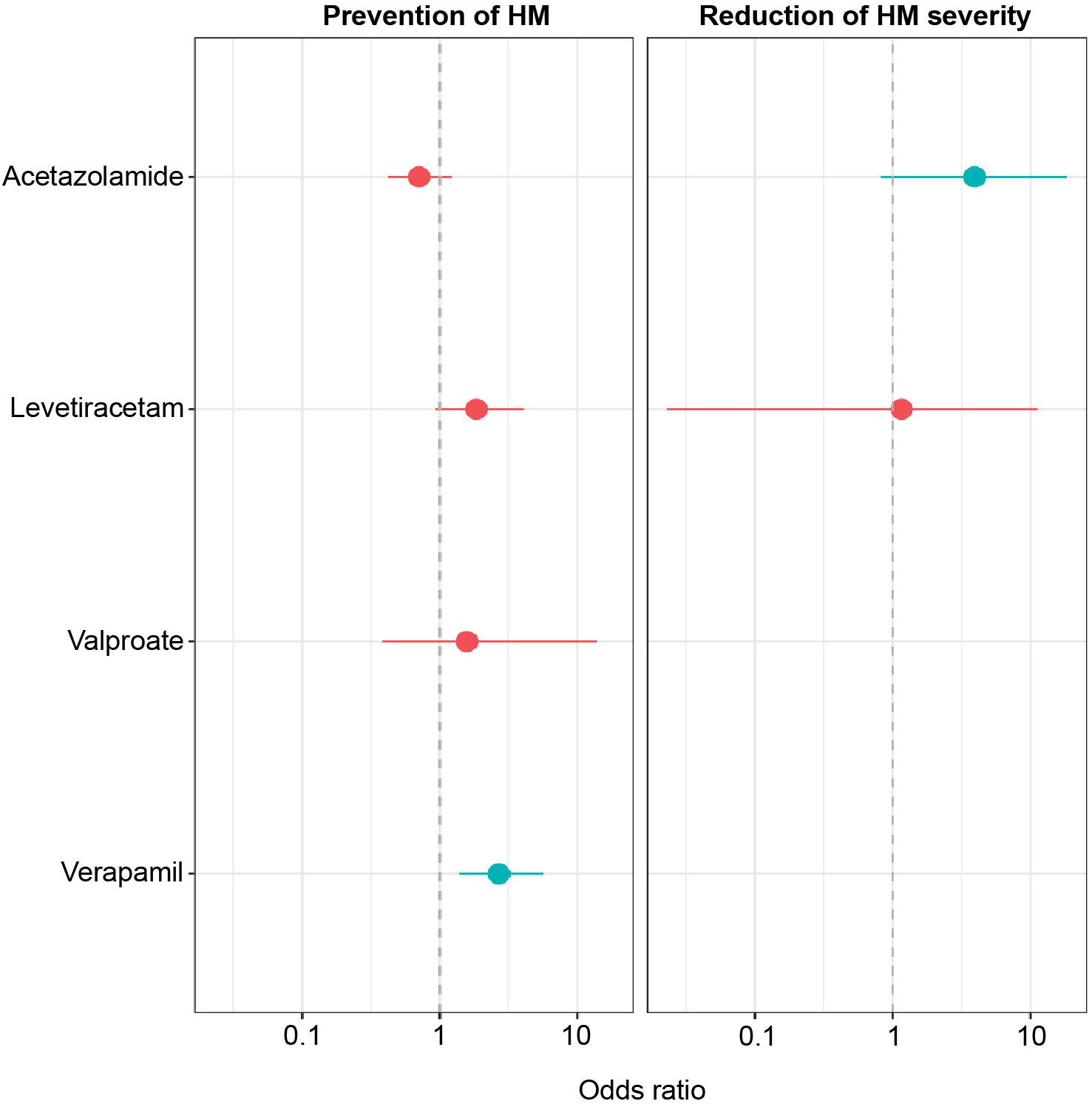
Distribution of prescriptions for preventative treatments for hemiplegic migraine, including anti-seizure medications. The frequency of medicine corresponds to the percentage of individuals on the specific medication at the age listed on the x-axis. Of note, this does not include rescue medications.

Using a previously established method to assess relative efficacy of treatments on reconstructed longitudinal data^28^, we assessed the impact on HM frequency and severity for treatments used in at least three individuals: levetiracetam, acetazolamide, verapamil, and valproate (**Figure 4**). In terms of the HM severity reduction, after correction for multiple testing, acetazolamide (*n* = 5, *p* = 0.045, OR 3.95, CI 0.82-18.37) showed only a nominally significant result, while levetiracetam (*n* = 3, *p* = 1, OR 1.16, CI 0.02-11.33) showed no significant results. On the other hand, the use of verapamil was significantly associated with the prevention of HM following the correction for multiple testing (*n* = 3, *p* = 0.002, OR 2.68, CI 1.39-5.67). The remainder of the medications did not have significant associations with the prevention of HM: levetiracetam (*n* = 6, *p* = 0.09, OR 1.84, CI 0.93, 4.07), acetazolamide (*n* = 5, p = 0.21, OR 0.71, CI 0.42-1.23), valproate (*n* = 3, *p* = 0.76, OR 1.57, CI 0.39-13.80).

**Figure 4.**
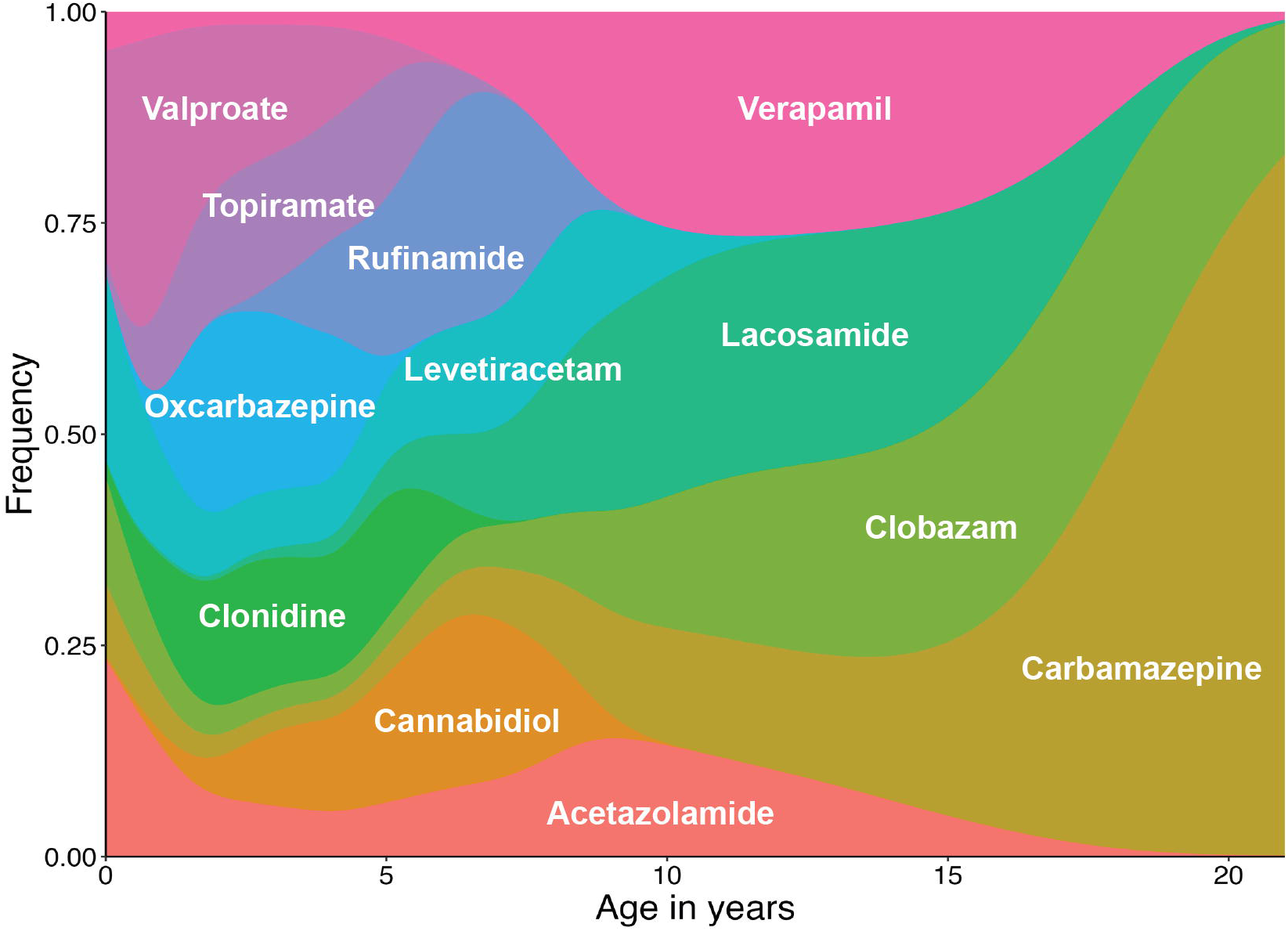
Comparative usefulness of preventative therapeutics for hemiplegic migraine. Left to right: Prevention of hemiplegic migraine (HM) events; Reduction of HM severity. Blue indicates nominally statistical significance. For each individual, the frequency of events was compared to the month prior to determine if there was a decrease in frequency of HM events (left graph) or decrease in severity of HM events (right graph). OR = 1 means no change. OR > 1 (on left graph) indicates less frequency of HM events. OR > 1 (on right graph) indicates HM events with less severity. Only medications used in 3 or more individuals were included.

## Discussion

Here, we examine the longitudinal trajectory of hemiplegic migraine in children with *CACNA1A*-related disorders. Our study highlights the unpredictability and heterogeneity of *CACNA1A*-related disorders, which include hemiplegic migraine episodes of varying severity, epilepsy, and a range of neurodevelopmental features. Neuroimaging findings including brain atrophy were a common finding in our cohort.

We identified several notable findings with regards to the trajectory and severity of hemiplegic migraine events. First, events found in our cohort reflect the severe end of the spectrum reported in the literature: 25% of all HM events were severe, lasting greater than 3 days or involving cerebral edema, and the majority of the cohort (73%) had at least one severe HM in their lifetime. No clear patterns emerged regarding the severity or frequency of HM events either in individual study participants or in the cohort as a whole. These findings suggest that although the majority of HM episodes will not be life-threatening, most individuals will have at least one potentially life-threatening event in their lifetime. Given the unpredictability of HM episodes, a lack of prior severe HM events does not exclude the possibility of a future severe event. Finally, with first severe HM episodes observed as late as age 13, there does not appear to be a clear age at which an individual outgrows their risk for severe HM. Given that the full severity of an HM event may not be recognized immediately, all HM events in individuals with *CACNA1A*-related disorders should be treated as potentially life-threatening.

We also found that the majority of our study population had significant comorbid features, including brain atrophy, neurodevelopmental traits, and epilepsy. Brain atrophy was prevalent in the cohort at 40% with an average age of onset of 5 years, including both cerebral and cerebellar atrophy. Even though various degrees of cerebellar atrophy are a known feature of *CACNA1A*-related disorders, the early onset and involvement of supratentorial structures is out of proportion to what is known about *CACNA1A*-related disorders. This finding emphasizes the overall severity of *CACNA1A*-related disorders with HM in children. Global developmental delay (87.7%), autism spectrum disorder (53.3%), and communication deficits (53.3%) were common in our cohort and only a single individual was neurotypical. In addition, over 50% of the cohort had epilepsy with half those individuals having refractory seizures. This wide spectrum of comorbid features and severities highlights the complexity of *CACNA1A*-related disorders even outside of HM events and has significant implications for clinical management.

A surprising finding in our study was the presence of suspected episodes of encephalitis in 26.7% of individuals prior to their first hemiplegic migraine event. In retrospect, these early events, which included encephalopathy with status epilepticus, may reflect the same pathophysiology as HM events and may be precursors of HM. It is therefore important to consider *CACNA1A-*related HM in individuals with unusual infantile presentations of suspected encephalitis that do not meet diagnostic criteria for infection or inflammation, especially in children with comorbid developmental differences or epilepsy.

In addition, we identified mild head trauma for at least one episode in 26.7% of individuals, even though this trigger was only present in a minority of overall events. Mild head trauma is a well-known trigger for HM^4,10,25^ and our finding emphasizes while most individuals have at least a single event provoked by mild head trauma, most events did not have an identifiable trigger. We found limited evidence for the benefit of preventative medications, both with regard to the frequency and severity of HM events. This highlights the clinical complexity of *CACNA1A*-related HM.

## Conclusion

In summary, our study examines the longitudinal trajectory of *CACNA1A*-related HM in children, an understudied episodic neurological event with significant morbidity in affected individuals. Our study emphasizes the unpredictable nature of HM with risk of future events in affected individuals that may present as severe, resulting in life-threatening consequences that require immediate intervention. Triggers are largely unknown, with mild head trauma as an inconsistent trigger. Accordingly, in our clinical practice, we recommend that individuals with a *CACNA1A* variant that has potential to cause HM be provided an action plan for HM and guidance to treat all HM events as if they could be severe. Furthermore, neurodevelopmental features and epilepsy are very common comorbid features that add complexity to management and ultimate prognosis. In summary, our study provides insights into the natural history of *CACNA1A-*related HM that can serve as groundwork for further study of genotype-phenotype relationships, treatment efficacy, triggers, and delineation of the natural history through puberty into adulthood.

## Acknowledgments

We want to thank the patients and families with *CACNA1A*-related hemiplegic migraine for participating in this research study as well as the CACNA1A Foundation for their support of this investigation. Research funded by R01-NS127830.

## Data availability statement

Data and computer code available via github on request.

## Funding statement

See acknowledgements; research funded by R01-NS127830.

## Conflict of interest disclosure

None of the authors has any conflict of interest to disclose.

## Ethics approval statement

We confirm that we have read the Journal’s position on issues involved in ethical publication and affirm that this report is consistent with those guidelines.

## Patient consent statement

Legal guardians of participants gave verbal or written informed consent. Study approved by CHOP IRB 15-012226

## Author contributions

- **Donna Schaare**: conceptualization (equal), data curation (lead), formal analysis (supporting), investigation (lead), writing – original draft, writing – review and editing (equal)
- **Laina Lusk**: conceptualization (equal), data curation (supporting), investigation (supporting), methodology (supporting), writing – review and editing (equal)
- **Alexis Karlin**: conceptualization (equal), investigation (supporting), writing – review and editing (equal)
- **Michael C. Kaufman**: data curation (supporting), formal analysis (lead), visualization (lead), writing – review and editing (equal)
- **Jan Magielski**: data curation (supporting), formal analysis (supporting), visualization (supporting), writing – review and editing (equal)
- **Sara M. Sarasua**: writing – review and editing (equal)
- **Kendra Allison**: writing – review and editing (equal)
- **Luigi Boccuto**: supervision (equal), writing – review and editing (equal)
- **Ingo Helbig**: conceptualization (equal), funding acquisition, methodology (lead), supervision (equal), writing – review and editing (equal)

## Statements

- **Conflict of interest disclosure**: None of the authors has any conflict of interest to disclose.
- **Ethics approval statement:** We confirm that we have read the Journal’s position on issues involved in ethical publication and affirm that this report is consistent with those guidelines.
- **Patient consent statement:** Patients consented according to CHOP IRB 15-012226.

## Notes

### Competing Interest Statement

The authors have declared no competing interest.

### Funding Statement

The study was funded by R01-NS127830.

### Author Declarations

IRB of Children's Hospital of Philadelphia gave ethical approval for this work

