## Supplementary figures and images for "A Longitudinal Exploration of *CACNA1A*-related Hemiplegic Migraine in Children"

### Figure S1

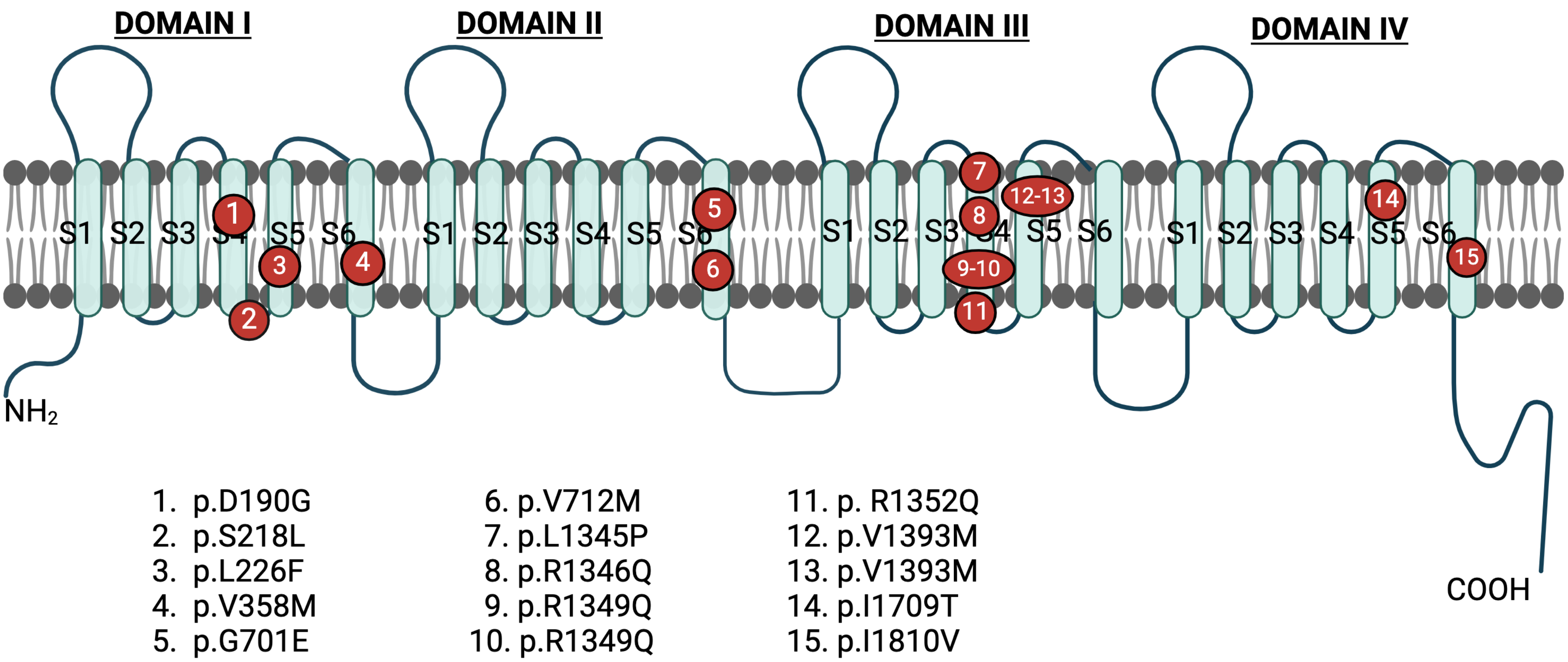

### Figure S2

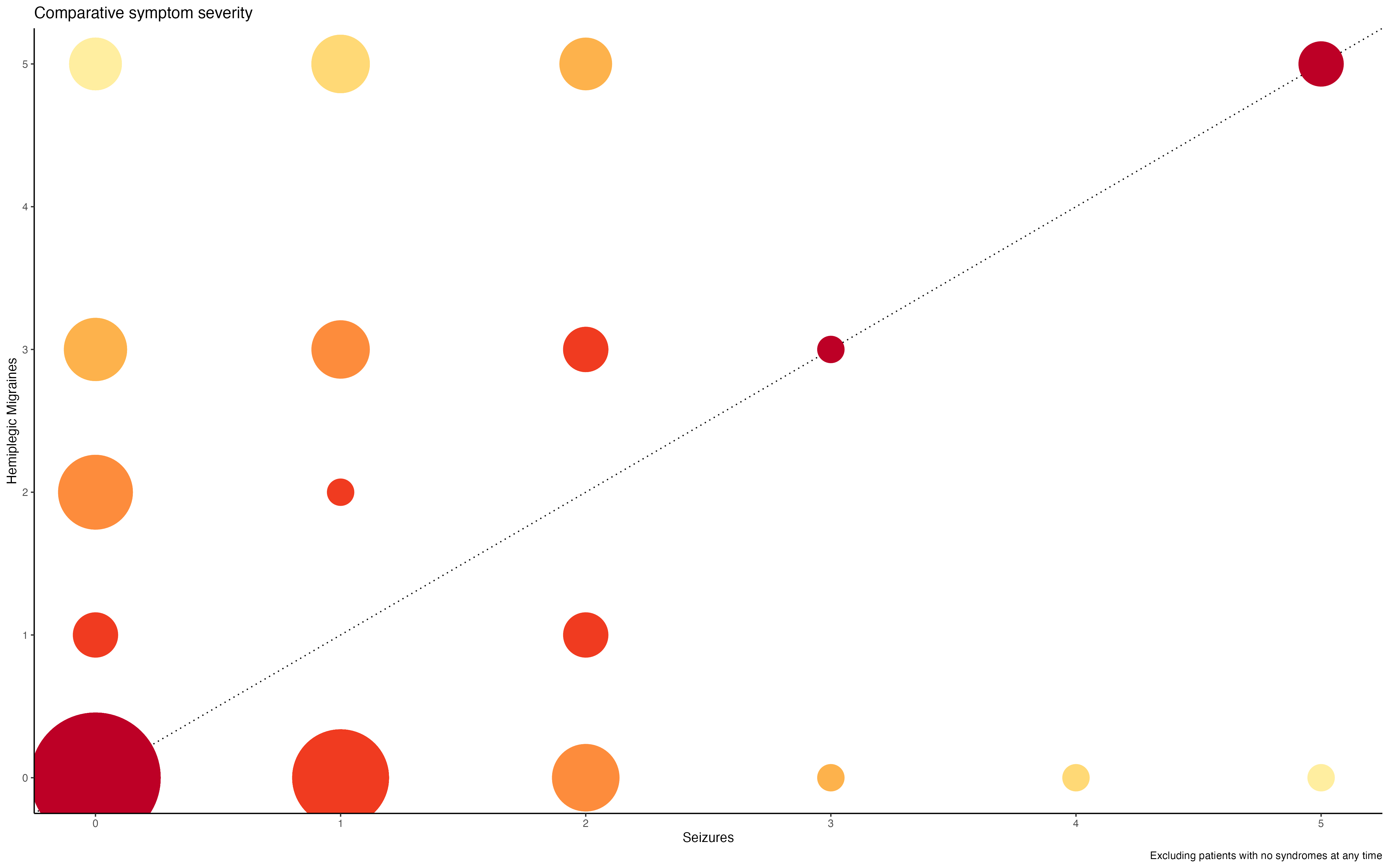
